# Analysis of 472,688 severe cases of COVID-19 in Brazil showed lower mortality in those vaccinated against influenza

**DOI:** 10.1101/2021.05.11.21257053

**Authors:** Daniele Melo Sardinha, Diana da Costa Lobato, Ana Lúcia da Silva Ferreira, Karla Valéria Batista Lima, Ricardo José de Paula Souza e Guimarães, Luana Nepomuceno Gondim Costa Lima

**Affiliations:** Post-graduate Program in Epidemiology and Health Surveillance of the Instituto Evandro Chagas (PPGEVS/IEC). Ananindeua, Pará, Brazil; State Coordinator of the Reference Center for Special Immunobiologicals from the Pará State Public Health Department (CRIE/SESPA). Belém, Pará, Brazil; Post-Graduation Program in Parasite Biology in the Amazon, University of state of Pará, Instituto Evandro Chagas (PPGBPA/UEPA/IEC). Belém, Pará, Brazil

**Keywords:** COVID-19, Pandemic, Brazil, Influenza Vaccine

## Abstract

**Objective:** To analyze the severe cases of COVID-19 in Brazil in 2020 and compare those vaccinated and unvaccinated against influenza in invasive ventilation, admission in Intensive Care Unit (ICU) and deaths.

**Method:** Cross-sectional study with public data from the OpenDataSUS platform, regarding confirmed severe cases for COVID-19 in Brazil in the year 2020. Data were analyzed by SPSS, from the chi-square test of independence and binary logistic regression.

**Results:** The population was 472,688 cases and 177,640 deaths, with a lethality of 37.58% in severe cases. The test of independence was highly significant in vaccinated survivors (<0.0001), and regression showed an almost twofold odds ratio for invasive ventilation, ICU admission, and death in unvaccinated cases.

**Conclusion:** We recommend mass influenza vaccination as an adjuvant in combating the COVID-19 pandemic in Brazil.

## 1. Introduction

The COVID-19 in Brazil represents a serious global public health problem, since one year after the pandemic, the country faces its worst scenario, with a record number of cases and deaths in the first three months of 2021 and a collapse in health services in some states, such as Amazonas, which faced oxygen shortages and thousands of deaths caused by the lack of the medicinal gas and also by the emergence of a new variant of SARS-CoV-2 [1, 2]. On 04/01/2021 Brazil has 12,839,844 confirmed cases and 325,284 deaths, of which 66,868 were in the month of March 2021 alone, surpassing July 2020, which was the worst-case scenario in 2020 [3].

Recently hypotheses have been raised that the trivalent influenza vaccine may be associated with lower severity and mortality in COVID-19. Some authors hypothesize that the association lies in sustained immunity, which is characterized by the recent stimulation of immune system responses to viral infections, which potentiated a better immune system response because it remains active through the toll-like receptor that is important for binding single-stranded RNA viruses, such as SARS-CoV-2. As well as cross-immunity, due to the structural similarities of the agents, such as the hemagglutinin protein [4, 5]. In this context, we wondered if the influenza vaccine is associated with lower mortality in COVID-19? Being the aim of the research to analyze the severe cases of COVID-19 in Brazil in 2020 and compare vaccinated and unvaccinated patients against influenza, in invasive ventilation, admission in Intensive Care Unit (ICU) and deaths.

## 2. Material and Method

A cross-sectional study was conducted and all severe cases and deaths of COVID-19 in Brazil were analyzed for the period from 03/01/2020 to 12/12/2020 from the database available on the public OpenDataSUS platform, which is data from the Influenza Epidemiological Surveillance Information System (SIVEP-Gripe) that reports and investigates hospitalized cases and deaths from SARS for all etiologies.

Only cases confirmed for COVID-19, with completed evolution, were included. Cases of deaths from other causes and blank progression were excluded. All confirmed cases for COVID-19 reported in 2020 with completed progression to cure or death from COVID-19 were included.

We performed the chi-square test of independence by 2 x 2 contingency table comparing vaccines between survivors and deaths and three binary logistic regression models, each with the dependent variable death, ICU admission and invasive ventilation, and in all three models the independent variable was not vaccinated against influenza. We used the Statistical Package for the Social Sciences (SPSS) software version 25.0.

The data of this study were publicly available, not containing personal data of patients such as name, address and telephone contact, presenting no risk to research participants, besides being exempt from ethical opinion. This study is carried out by Law No. 12,527 of 18/11/2011 (Access to Information Law) [6].

## 3. Results

The study population corresponded to 472,688 cases of COVID-19 Severe Acute Respiratory Syndrome (SARS), of which 177,640 died, with a mortality rate in severe cases of 37.58%. In Table 1, the test (X2 281,259) was highly significant for vaccinated survivors (<0.001), showing that influenza vaccine is a protective factor in survivors.

**Table 1.** Influenza vaccinees compared to survivors and deaths in the 472,688 in cases of SARS by COVID-19 in Brazil in 2020.

| Variable | Total<br>(472,668) | % | Survivors<br>(295,028) | % | Deaths<br>(177,640) | % | *X<sup>2</sup> | p-value<br>survivors<br>v.s. deaths |
| --- | --- | --- | --- | --- | --- | --- | --- | --- |
| Vaccinated<br>against<br>influenza | 62,771 | 13.28 | 41,955 | 14.22 | 20,816 | 11.71 | 281,259 | <0.001 |

Source: Ministry of Health, Sivep-Gripe/OpenDataSUS, 2020. \*chi-square.

The binary regression was significant in all three models (<0.001), regarding the outcome of invasive ventilation, the model showed the odds ratio (OR 1.451/CI 1.417-1.486). In ICU admission (OR 1.328/CI 1.303-1356). And in deaths (OR 1.249/CI 1.227-1.271). Thus, all outcomes analyzed were significant in unvaccinated cases, showing that not being vaccinated against influenza has higher chances of ICU admission, invasive ventilation and death, when compared to vaccinated cases (table 2).

**Table 2.** Association of ICU inpatients, invasive ventilation, and death v.s. not vaccinated against influenza, in cases of SARS by COVID-19 in Brazil in 2020.

| Not vaccinated against influenza | Test | df | p-value | Odds ratio | 95% C.I* for Odds ratio |  |
| --- | --- | --- | --- | --- | --- | --- |
|  |  |  |  |  | Lower | Upper |
| Invasive ventilation | 922.680 | 1 | < 0.001 | 1.451 | 1.417 | 1.486 |
| Constant | 25081.133 | 1 | < 0.001 | 0.160 |  |  |
| ICU admission | 891.416 | 1 | <0.001 | 1.328 | 1.303 | 1.356 |
| Constant | 11449.833 | 1 | < 0.001 | 0.386 |  |  |
| Death | 601.280 | 1 | < 0.001 | 1.249 | 1.227 | 1.271 |
| Constant | 6834.456 | 1 | < 0.001 | 0.496 |  |  |

Source: Ministry of Health, Sivep-Gripe/OpenDataSUS, 2020. \*Confidence interval.

## 4. Discussion

Some studies have also identified the association of influenza vaccine in reducing COVID-19 mortality. The research by Wilcox, Islam, and Dambha-Miller [7] was associated with lower hospitalization and all-cause mortality (adjusted OR: 0.85, 95% CI 0.75 to 0.97, p = 0.02), and 24% reduced odds of all-cause mortality (adjusted OR: 0.76, 95% CI 0.64 to 0.90) with 15% and 24% lower odds of these outcomes in severe cases of COVID-19. A study in Brazil showed that those vaccinated against influenza were 8% less likely to be admitted to the intensive care unit, 18% less likely to require mechanical ventilation, and 17% less likely to die[8]. A study in Italy analyzed data from vaccinated and unvaccinated elderly over 65 years of age, and showed moderate to strong negative correlation (r = -.5874, n = 21, P = .0051), i.e., when there were higher rates of influenza vaccination, fewer deaths from COVID-19 occurred. [9]. Research by Yang et al[10] showed that COVID-19 patients who had not been vaccinated against influenza in the past year had a 2.44 (95% CI, 1.68, 3.61) higher chance of hospitalization and 3.29 (95% CI, 1.18, 13.77) higher chance of ICU admission when compared to those vaccinated.

In this context, some hypotheses exist about other vaccines as a protective factor in mortality by COVID-19, as for example the BCG vaccine, the author highlights that BCG vaccination administered at birth can exert heterologous immune effects to increase protection against unrelated pathogens, besides protection against tuberculosis [11]. For O’Neill and Netea [12], BCG vaccine has the potential to reprogram innate immunity, protecting against respiratory infections, because it is an attenuated vaccine administered as soon as the individual is born. Randomized studies have shown a 50% reduction in mortality in children vaccinated with BCG; thus, since most of the causes of death in children are respiratory infections, BCG vaccine has been associated with a reduction in deaths from all causes in children [13]. Thus, based on these hypotheses raised, our results may be associated with BCG protection, however our database for the analysis of this research did not have this information about BCG vaccination, only influenza, so the limitation of the study stands out.

Thus, our results from analysis of all severe cases of COVID-19 in 2020 in Brazil also associated that influenza vaccine reduces invasive ventilation, ICU admission and death, corroborated by some studies in the literature.

## 5. Conclusion

We evidenced that the influenza vaccine is associated with a lower chance of ICU admission, invasive ventilation and death, although the immunological mechanisms responsible are still unclear in the literature, they are still hypotheses, however, the results of this research and other studies confirm this association of vaccination with better outcome in severe cases of COVID-19.

We recommend that influenza vaccination be used as a secondary medicine to combat the pandemic of COVID-19, including individuals >6 months of age, as well as to ensure high vaccination coverage in the elderly population. To achieve this goal, influenza vaccine should be included in routine as an annual dose, regardless of specific influenza campaigns for the elderly and priority groups, as it will increase access and coverage.

## Data Availability

opendatasus

## Footnotes

** Special description of the title. (dispensable)

